## Supplementary information for "The United Kingdom Research study into Ethnicity And COVID-19 outcomes in Healthcare workers (UK-REACH): Protocol for a prospective longitudinal cohort study of healthcare and ancillary workers in UK healthcare settings"

List of Members of UK-REACH Stakeholder Group (STAG) as at January 2021

1. General Medical Council (GMC)

2. Nursing and Midwifery Council (NMC)

3. General Dental Council (GDC)

4. General Pharmaceutical Council (GPC)

5. Royal College of Psychiatrists (RCPsych)

6. Royal College of Obstetricians and Gynaecologists (RCOG)

7. Royal College of Midwives (RCM)

8. NHS Confederation

9. British Association of Physicians of Indian Origin (BAPIO)

10. Sudan Doctors’ Union –UK Branch

11. Association of Pakistani Physicians of Northern Europe (APPNE)

12. South Asian Health Foundation (SAHF)

13. Health Education England (HEE)

14. General Optical Council (GOC)

15. Filipino Nurses Association UK (FNAUK)

16. Pharmaceutical Society of Northern Ireland (PSNI)

17. Health and Care Professions Council (HCPC)

18. NHS England & Improvement

19. British Medical Association (BMA)

20. Medical Association of Nigerians Across Great Britain (MANSAG)

List of Members of UK-REACH Professional Expert Panel (PEP) as at January 2021

Susie Lagrata (co-Chair), Nurse.

Padmasayee Papineni (co-Chair), Doctor.

Sandra Kazembe, Nurse.

Tatiana Monteiro, Domestic worker.

Juliette Mutumiyana, Doctor.

Satheesh Mathew, Doctor.

Amir Burney, Doctor.

Ahmed Hashim, Doctor.

Tiffanie Harrison, Nurse.
